# Intraoral Ultrasound for Detection of Alveolar Bone Changes Following Periodontal Surgery: A Prospective Validity and Precision Study

**DOI:** 10.64898/2026.06.29.26356850

**Authors:** Mirali Pandya, Bryant Tran, Mohammadreza Amjadian, Sabrina Alterman, Harrison Chang, Youyoung Min, Suhel Khan, Jesse Jokerst, Casey Chen

## Abstract

**Background:** Alveolar bone assessment in periodontal practice relies on radiography and clinical probing, both of which have well-documented limitations in precision. Intraoral high-frequency ultrasonography (US) offers a radiation-free alternative with potential for sub-millimeter resolution, the validity and precision for detecting minute osseous changes have not been established. The purpose of this study was to evaluate the concurrent validity and measurement precision of intraoral US for detecting alveolar bone-level changes in patients undergoing crown lengthening and osseous surgery, thereby enabling its translation to monitor osseous changes in patients with periodontitis.

**Methods:** Ten patients (28 tooth sites) undergoing crown lengthening or osseous surgery at a USC Advanced Grad Perio clinic were enrolled in this prospective observational study. Distance from the cementoenamel junction (CEJ) to the Alveolar bone crest (ABC) was measured at pre- and post-operative time points using a 40 MHz handheld intraoral US transducer and, intraoperatively, by standardized clinical photography. Agreement was assessed by Pearson correlation and Bland-Altman analysis. Measurement precision was quantified using the standard error of measurement (SEM) and minimum detectable change (MDC).

**Results:** Preoperative agreement between methods was excellent (r = 0.977; Bland-Altman bias = −0.009 mm; 95% limits of agreement [LoA]: ±0.40 mm). Post-operative correlation remained strong (r = 0.912; bias = 0.123 mm; LoA: −0.85 to +1.10 mm). Both methods detected statistically significant post-surgical increases in the ABC-to-CEJ distance (p < 0.001), as anticipated. US demonstrated substantially superior precision: preoperative SEM 0.058 mm with US versus 0.128 mm clinically, yielding MDC values of 0.160 mm (US) versus 0.354 mm (clinical), providing a 2.2-fold precision advantage.

**Conclusions:** Intraoral US demonstrated strong concurrent validity with clinical photography and a reproducible precision advantage in detecting alveolar bone-level changes in patients with periodontitis. These findings support its clinical utility as a radiation-free, high-sensitivity bone monitoring tool. Larger longitudinal studies with CBCT validation are warranted.

## INTRODUCTION

Periodontitis is one of the most common chronic inflammatory diseases in the United States and worldwide, affecting the gingiva, periodontal ligament, cementum, and alveolar bone, and representing a leading cause of tooth loss in adults.[1, 2] Periodontal disease is characterized by progressive destruction of alveolar bone and is irreversible once it surpasses a critical threshold of transitioning from gingivitis to loss of interproximal clinical attachment and bone.[3] Early and accurate detection of bone-level changes is therefore central to effective disease management as it guides our diagnosis, informs appropriate treatment planning, and provides the objective benchmarks against which the risk of progression and therapeutic outcomes are measured.

Periapical radiography has served as the clinical standard for assessing the alveolar bone in periodontal practice for decades. As a two-dimensional projection modality, radiographs are inherently insensitive to buccal and lingual bone changes, prone to superimposition artifacts, and subject to variable angulation and exposure geometry that are inevitably encountered in routine clinical use.[2, 4] There are several limitations in using radiography to detect bone level changes. The radiographic bone loss becomes detectable only after approximately 30–50% of the cortical alveolar bone has already been lost.[5] Another limitation is the relatively large system error of plain-film radiographic analysis, and any changes in bone level of 0.87 mm or less by radiographs cannot be used to diagnose bone loss over time[6]. These limitations place the clinician well behind the biological curve in identifying early disease.[7] Finally, repeated ionizing radiation exposures required for radiographic monitoring of alveolar bone levels, though modest at the level of an individual examination, also remain a concern for patients requiring serial monitoring at frequent intervals.

Against this background, ultrasonography (US) has attracted growing attention as an alternative imaging modality for periodontal assessment. High-frequency intraoral US transducers (typically 10–40 MHz) are capable of resolving soft and hard tissue interfaces at sub-millimeter precision without ionizing radiation, without requiring inter-appointment intervals, and in a format compatible with routine chairside use.[8, 9] A series of validation studies over the past decade have examined its ability to measure gingival thickness, alveolar bone level, and the distance from the alveolar bone crest to the cementoenamel junction (ABC-CEJ). These parameters are of significant clinical importance in periodontal diagnosis and monitoring.[10–12] Figueredo et al., in a 2023 reliability study using a 20 MHz handheld intraoral transducer, demonstrated excellent intra-rater reliability for ABC-CEJ measurements (ICC 0.940) across 64 patients, while Moore et al. reported that a 40 MHz system achieved equivalent diagnostic capacity to conventional probing for key periodontal parameters in both healthy and diseased subjects.[13, 14]

Our group’s latest proof-of-concept article by Abdelrahim et al.[12] demonstrated in an ex vivo porcine model that the minimum detectable increment in bone loss using a 40 MHz high-frequency US transducer was approximately 138 µm.[12] Despite this accumulating body of evidence for baseline US validity, far less is known about its performance in a longitudinal or interventional context specifically, and whether US can reliably detect and quantify the discrete osseous changes produced by periodontitis. Crown lengthening and osseous surgeries provide a controlled and clinically relevant model to address this question. The surgical procedure involves deliberate apical displacement of the alveolar crest through a combination of ostectomy and osteoplasty, producing measurable changes in bone level that can be assessed pre- and post-intervention.[15, 16] It therefore represents an opportunity to evaluate the concurrent validity of US against an established clinical reference and its sensitivity to real, surgically induced osseous change, which could prove to be a property that is ultimately more important for longitudinal monitoring than static agreement alone.

The purpose of our study was to evaluate the validity and precision of intraoral ultrasound for measuring alveolar bone-level change in patients undergoing periodontal crown lengthening and osseous surgery, using standardized clinical photography as a reference. Specific objectives were to: (1) characterize preoperative and post-operative agreement between US and clinical measurements, (2) compare the intrinsic measurement precision of the two modalities through calculation of the standard error of measurement (SEM) and minimum detectable change (MDC).

## MATERIALS AND METHODS

### Patient Selection and Ethical Approval

Ten adult patients scheduled to undergo crown lengthening or osseous surgery at the Herman Ostrow School of Dentistry Advanced Graduate Periodontics Clinic were enrolled for our prospective study. The study was conducted in accordance with the protocol approved by the institutional review board for human subjects research at USC (# HS-19-00163). All patients were provided with written informed consent before enrollment. Data collection and analysis were performed by a trained operator with experience in intraoral ultrasound image acquisition and Graduate Periodontics residents performed the surgery at USC.

Patients were eligible for participation if they required surgical crown lengthening or probing depth reduction at one or more tooth sites, were systemically healthy or medically stable, and were nonsmokers or former smokers with no active smoking within the preceding 12 months. Exclusion criteria included pregnancy, use of medications known to affect bone metabolism, presence of acute periodontal infection at the study site, or inability to attend the post-operative assessment. Surgical indications included subgingival caries, crown or root fracture requiring ferrule preparation, inadequate clinical crown height for restorative purposes, and deep periodontal probing depths with osseous crater defects. A total of 28 tooth sites across the 10 subjects were included in the final analysis, following exclusion of sites with unacceptable image quality on US or clinical acquisition.

### Surgical Protocol

All surgical procedures were performed by a periodontics resident under the supervision of a board-certified and experienced periodontist. Following intrasulcular incisions and full-thickness mucoperiosteal flap reflection, the osseous architecture was visualized directly. Ostectomy and osteoplasty were performed using rotary instrumentation and hand instruments to achieve a minimum of 3 mm of distance between the ferrule and the alveolar bone crest, with careful attention to biologic width requirements for functional crown lengthening surgery.[17] For the osseous surgery, the gingival collar on the buccal and/or palatal aspects was removed based on clinical assessment, along with reshaping the alveolar bone to facilitate flap adaptation and eliminate deep periodontal probing depths. Flaps were repositioned slightly apical to the osseous crest and secured with interrupted or continuous sutures. Patients were placed on a standardized post-operative regimen including analgesics, antibiotics, and dietary guidance. Post-operative assessments were scheduled between two to six weeks following surgery, consistent with the period required for initial osseous maturation and soft tissue stabilization.[15]

### Clinical Measurement Protocol

The distance between the CEJ and alveolar bone crest was measured clinically from standardized intraoral digital photographs obtained pre- and post-osseous adjustment after flap reflection. A calibrated periodontal probe was positioned at the facial and palatal/lingual midpoints of each tooth included in the study to provide a scale reference within each image. The distance from the CEJ to the alveolar bone crest was measured digitally in each photograph using image analysis software (ImageJ, National Institutes of Health, Bethesda, USA), with the William’s probe used to convert pixel measurements to millimeters. To correct any photographic angulation, distances between bone crests and CEJs were quantified from intraoral images using a custom image analysis tool (Periodontal Bone-to-CEJ Measurement Tool). This software was written in Python using the Matplotlib/Tkinter libraries. Measurement was done using a sequential six-step process. Two calibration points of known distance apart on the periodontal probe were detected first in order to obtain the pixels/millimeter scale. Bone crests and CEJs were detected next by point clicks, resulting in a raw Euclidean distance in millimeters. In-plane angulation correction calculated the acute angle (θ) between the measurement vector (bone crest to CEJ line) and the long axis of the probe (two points clicked along the probe shaft) and divided the raw measurement by cos(θ) due to the measurement vector not being parallel to the probe shaft. Out-of-plane camera tilt correction calculated the tilt angle (α) as the ratio of apparent probe shaft diameter in pixels scaled to millimeters using previously obtained scale and the actual probe shaft diameter (1.0 mm); the in-plane corrected measurement was then divided by cos(α) to account for foreshortening due to camera angulation out of the plane of the image. Final distance was calculated as follows: final distance (mm) = raw_mm ÷ cos(θ) ÷ cos(α). All clinical measurements were performed three times by a single examiner blinded to the corresponding US findings using ImageJ software.

### Ultrasound Measurement Protocol

Intraoral ultrasound imaging was performed using a handheld 40 MHz high-frequency transducer positioned at the straight facial and palatal/ lingual aspect of each study tooth. The transducer was oriented perpendicular to the long axis of the tooth, and acoustic coupling was performed using a custom hydrogel sleeve containing a small amount of ultrasound transmission gel (Aquasonic® Clear Ultrasound Gel; Parker Laboratories, Inc., Fairfield, NJ, USA). The sleeve was covered by a transparent sheet (Tegaderm™ Pad Film Dressing; 3M Company, St. Paul, MN, USA. For each site, the transducer was positioned to identify the alveolar bone crest (ABC), cementoenamel junction (CEJ) and the gingival margin (GM). In cases with a temporary crown in which CEJ was no longer present, the border of the temporary crown served as the landmark instead of CEJ. Alveolar bone height was computed as the distance between the CEJ/ crown imaging echo and the bone crest echo, as identified on the resulting B-mode image. Image acquisition was performed by a separate trained operator at both timepoints, and landmark identification was performed by a different trained operator. All US measurements were performed three times with spaced intervals by a single examiner blinded to the corresponding clinical results..

### Statistical Analysis

All statistical analyses were performed by Python with scipy, pandas, numpy and matplotlib libraries. Descriptive statistics (mean, standard deviation) were calculated for alveolar bone level change at each timepoint and for each method. Pearson correlation coefficients were calculated to quantify the linear relationship between US and clinical measurements at each timepoint and for the magnitude of surgical change. Bland-Altman analysis was performed to assess systematic bias and 95% limits of agreement (LoA) between methods.[18] The standard error of measurement (SEM) was calculated as the mean of the intra-session standard deviation of three repeated measurements per site, and the minimum detectable change (MDC) was defined as 1.96 × √2 × SEM, representing the smallest change distinguishable from measurement error at the 95% confidence level.[19] A two-tailed p-value of <0.05 was considered statistically significant for all comparisons. The software used to correct probe angulation in clinical images was custom-built in Python.

## RESULTS

### Sample and Descriptive Statistics

Correlation and agreement between ultrasound and clinical alveolar bone change measurements, along with the distribution of surgically induced change, are presented in Figure 1. Preoperatively, US and clinical measurements demonstrated excellent linear agreement across the 28 evaluable tooth sites, with a Pearson correlation of r = 0.977 (p < 0.001) and a near-zero bias of −0.009 mm, indicating no systematic directional offset between the two modalities at baseline. The regression line closely followed the identity line across the full range of measured values, suggesting that the two methods were largely interchangeable for characterizing alveolar bone change. Postoperatively, the correlation between methods remained strong (r = 0.912, p < 0.001), though with a modestly increased bias of 0.123 mm and greater scatter at the upper range of alveolar bone level change (Panel B), consistent with the widened post-operative limits of agreement observed in the Bland-Altman analysis for the full cohort. The correlation between US-detected and clinically-detected surgical change was moderate-to-strong (r = 0.802, p < 0.001; Panel C), confirming that US reliably tracked the direction and relative magnitude of osseous displacement across individual sites.

**Figure 1.**
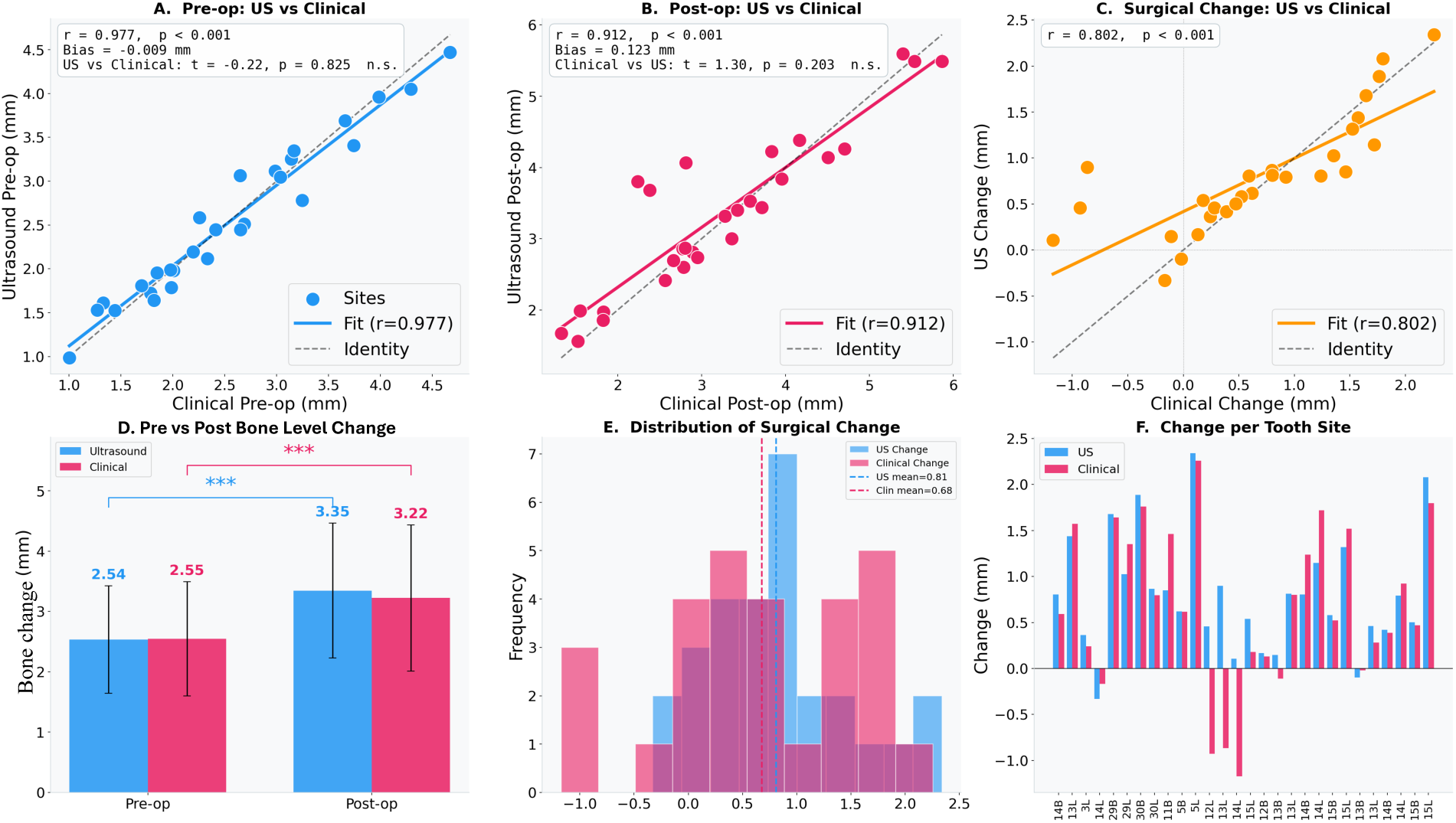
Correlation, descriptive statistics, and site-level distribution of ultrasound and clinical alveolar bone change measurements across pre-operative and post-operative timepoints. (A) Scatter plot of pre-operative ultrasound (US) versus clinical alveolar bone level change measurements across all evaluable tooth sites (n = 28). The regression line (solid blue) closely approximates the identity line (dashed), with excellent agreement (r = 0.977, p < 0.001, Bland-Altman bias = −0.009 mm). (B) Scatter plot of post-operative US versus clinical measurements. Correlation remained strong (r = 0.912, p < 0.001, bias = 0.123 mm), with modestly increased scatter at larger alveolar bone changes relative to the pre-operative comparison. (C) Scatter plot of US-detected versus clinically-detected magnitude of surgical change per site (r = 0.802, p < 0.001), demonstrating that US tracked the direction and relative magnitude of osseous change consistently with the clinical reference across individual sites. (D) Mean alveolar bone level change (mm ± SD) by modality at pre-operative and post-operative timepoints. Both US (2.54 → 3.35 mm) and clinical measurement (2.55 → 3.22 mm) detected a statistically significant post-surgical increase in alveolar bone level measurement (p < 0.001). Error bars represent one standard deviation. (E) Frequency distribution of surgical bone level change detected by US (blue) and clinical measurement (pink) across all sites. Dashed vertical lines indicate group means (US mean = 0.81 mm; Clinical mean = 0.68 mm). The majority of sites demonstrated positive change values consistent with apical displacement of the alveolar crest post surgery. (F) Site-level bar chart illustrating US (blue) and clinical (pink) change scores for each individual tooth site. Sites with negative or near-zero change values represent anatomical outliers or sites with minimal planned bone removal. US = ultrasound; CEJ = cementoenamel junction; ABC = alveolar bone crest.

Both modalities detected a statistically significant increase in the mean CEJ-to-ABC distance following surgery (Panel D). US-measured alveolar bone level change increased from a preoperative mean of 2.54 mm (SD ± 0.73) to a post-operative mean of 3.35 mm (SD ± 0.90). In comparison, clinical measurement yielded corresponding values of 2.55 mm (SD ± 0.72) and 3.22 mm (SD ± 0.89), reflecting mean surgical changes of 0.81 mm and 0.68 mm, respectively (p < 0.001 for both). The distribution of site-level surgical change is shown in Panel E. The majority of sites demonstrated positive change values of 0-1.5 mm, consistent with the planned degree of osseous resection across the cohort. Site-level variability in the magnitude of US-detected and clinically-detected change is further illustrated in Panel F, where directional concordance between modalities was maintained at most individual sites. A small number of sites showed near-zero or negative change values clinically, reflecting sites where either minimal bone amount was removed or the clinical photograph had an angulation that could not be corrected by the software, resulting in inaccurate measurement.

### Clinical data and ultrasound data comparison

A representative case illustrating the multimodal assessment approach is presented in Figure 2. Preoperatively, the gingival surface, CEJ, and ABC were each identifiable as discrete acoustic interfaces on B-mode ultrasound imaging, with contour delineation confirmed by the trained operator before measurement. The preoperative CEJ-to-ABC distance measured by ultrasound was 1.2 mm, compared to 1 mm by clinical photographic measurement. Postoperatively, the ABC echo demonstrates a clear apical displacement relative to its preoperative position, consistent with the ostectomy and osteoplasty. The post-operative CEJ-to-ABC distance was 3.5 mm by ultrasound and 4 mm clinically, yielding a surgically induced change of 2.3 mm and 3 mm, respectively, as detected by each modality. The direction and approximate magnitude of osseous change were concordant between methods, consistent with the moderate-to-strong post-operative correlation observed across the full cohort. Periapical radiographs obtained at each timepoint corroborated the expected mesiodistal interproximal bone-level changes, though buccal and palatal osseous displacement of #13, which was the primary target of the surgical procedure, was not fully appreciable on the two-dimensional radiographic projection. This underscores the dimensional limitation of conventional radiography relative to the direct buccal and palatal interrogation afforded by the intraoral ultrasound transducer. This case is representative of the pattern observed throughout the study, with high preoperative agreement between ultrasound and clinical measurements, maintaining directional concordance and moderately wider limits of agreement at the postoperative timepoint.

**Figure 2.**
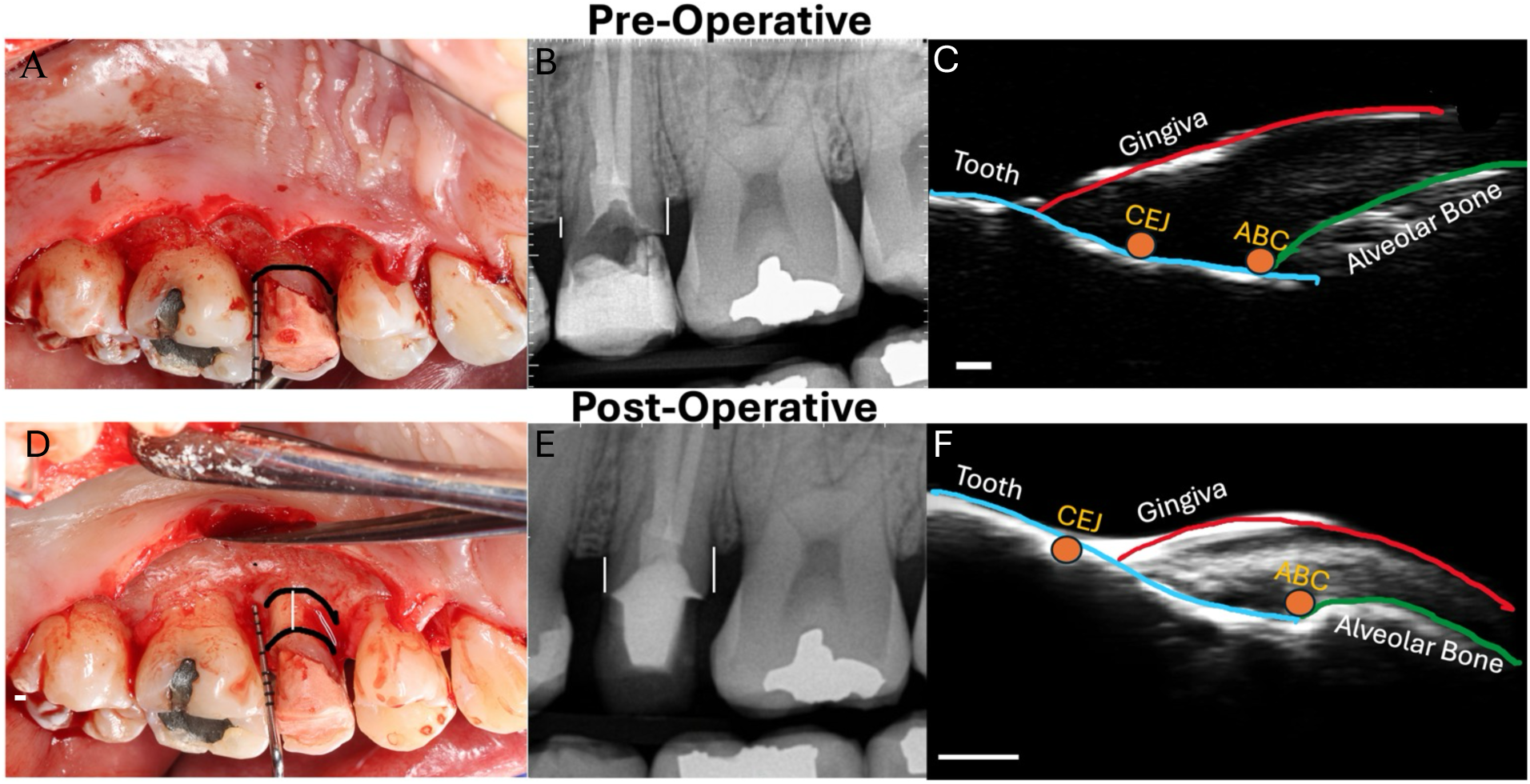
Multimodal pre-operative and post-operative assessment of alveolar bone-level change following functional crown lengthening for tooth #13. Each row represents a single timepoint: pre-operative (top panel A,B, C) and post-operative (bottom panel D, E, F). Left column shows intraoral clinical photograph with the palatal view obtained during direct osseous visualization following full-thickness mucoperiosteal flap reflection, serving as the photographic reference standard for bone level change measurement with a standardized periodontal probe for reference. Middle column: Corresponding periapical radiographs demonstrating mesiodistal alveolar bone architecture at each timepoint. Right column: B-mode intraoral ultrasound images with anatomical landmark annotation. The gingival surface (red contour), tooth-tissue interface (blue contour), and alveolar bone surface (green contour) are delineated. Yellow circles indicate the cementoenamel junction (CEJ) and alveolar bone crest (ABC), between which the primary outcome measurement showing CEJ-to-ABC distance that was calculated. Post-operative ultrasound demonstrates apical displacement of the ABC echo relative to the CEJ, consistent with surgically induced osseous change. White lines indicative of differences pre and post operatively. Scale bar = 1 mm for (C) and 2mm for (F).

### Bland Altman Analysis for Inter-method Agreements

Bland-Altman analyses are presented in Figure 3, organized to ask a distinct analytical question: did the measurements from ultrasound and clinical photography agree with one another at each timepoint.

**Figure 3.**
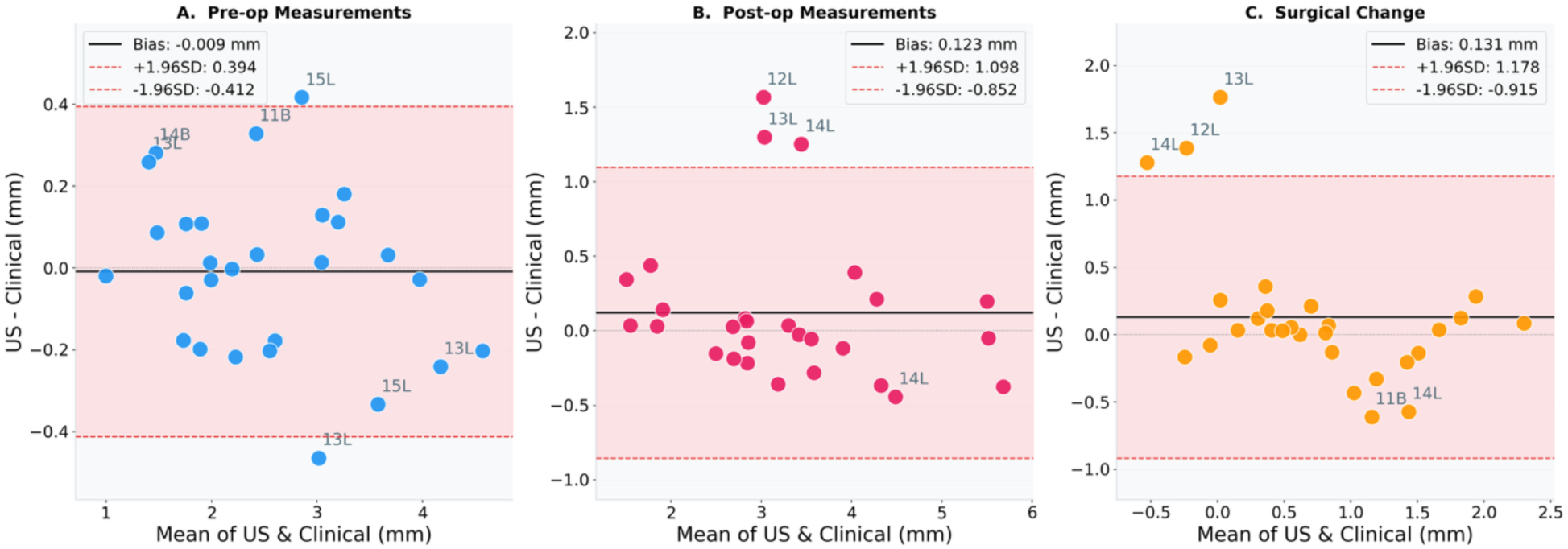
Bland-Altman analysis of between-method agreement for ultrasound and clinical alveolar bone level measurements. Between-method agreement comparing ultrasound and clinical photography measurements at three analytical stages. The y-axis represents the US minus clinical difference. (A) Pre-operative agreement (bias = −0.009 mm; 95% LoA: −0.412 to +0.394 mm). (B) Post-operative agreement (bias = +0.123 mm; 95% LoA: −0.852 to +1.098 mm). (C) Surgical change agreement, comparing the magnitude of change detected by each method (bias = +0.131 mm; 95% LoA: −0.915 to +1.178 mm). Labeled outlier sites (12L, 13L, 14L, 15L, 11B, 11B) represent tooth sites contributing disproportionately to inter-method disagreement and are discussed in the text. US = ultrasound; CEJ = cementoenamel junction; LoA = limits of agreement. Red dashed lines = 95% LoA; solid black line = mean bias.

Addressing the question, preoperatively, agreement between ultrasound and clinical photography was excellent (Panel A). The mean bias was −0.009 mm, statistically indistinguishable from zero, with 95% limits of agreement spanning −0.412 to +0.394 mm with a total spread of 0.806 mm. Data points were symmetrically distributed around the bias line across the full range of mean values, with no evidence of proportional bias or systematic directional offset between methods. Two sites (15L and 11B) approached or marginally exceeded the limits of agreement, but the overall pattern was one of tight inter-method concordance consistent with the preoperative Pearson correlation of r = 0.977. Postoperatively (Panel B), mean bias remained small at +0.123 mm. However, the limits of agreement widened substantially to −0.852 to +1.098 mm, representing a 2.4-fold increase in total span relative to the preoperative comparison (1.950 mm vs. 0.806 mm). This widening was not uniformly distributed across sites. Several tooth sites (12L, 13L, and 14L) appear as outliers above the upper limit of agreement. These outliers were from the same patient. For the surgical change comparison, the mean bias was +0.131 mm with 95% LoA of −0.915 to +1.178 mm (total span = 2.093 mm; Panel C).

### Standard Error of Measurement (SEM) and Minimum Detectable Change (MDC)

Measurement precision analysis is presented in Figure 4. Ultrasound demonstrated substantially lower SEM than clinical photography at both timepoints (Panel A). Preoperatively, US SEM was 0.058 mm compared to 0.128 mm for clinical photography, which represents a 2.2-fold difference. Postoperatively, this precision advantage was maintained, with the US SEM of 0.049 mm versus 0.096 mm clinically, representing a 1.96-fold difference. The corresponding MDC values, representing the smallest change at each site that can be distinguished from measurement noise at 95% confidence (Panel B). Preoperatively, the US MDC was 0.160 mm, compared with 0.354 mm for clinical photography. Postoperatively, MDC values were 0.136 mm (US) and 0.267 mm (clinical). Taken together, these figures confirm that ultrasound was approximately 2.2 times more sensitive than clinical photography for detecting true osseous change above the precision noise floor at the preoperative timepoint, and approximately 1.96 times more sensitive postoperatively. Panel C shows the precision thresholds as a function of the magnitude of the observed surgical change. The mean US-detected surgical change was 0.813 mm, and the mean clinically-detected change was 0.679 mm. Both methods substantially exceeded all four MDC thresholds and exceeded the 0.5 mm clinical significance threshold (orange dashed line based on clinical judgment). The observed surgical change exceeded the preoperative US MDC by a factor of 5.1 and the preoperative clinical MDC by a factor of 1.9, confirming that the osseous changes produced by crown lengthening and osseous surgery in this cohort were comfortably within the detection range of both modalities.

**Figure 4.**
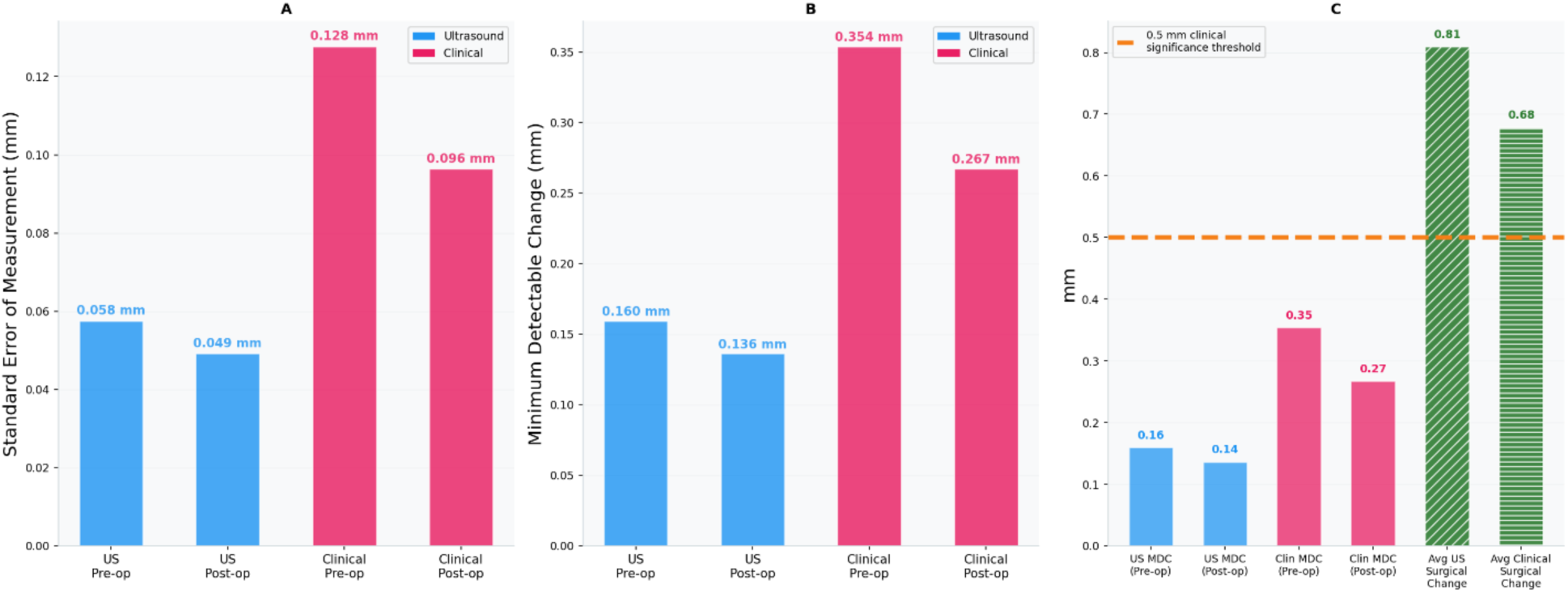
Minimum Detectable Change (MDC) analysis comparing measurement precision of ultrasound and clinical photography at pre-operative and post-operative timepoints, and contextualizing precision thresholds against the magnitude of observed surgical change. MDC represents the smallest change distinguishable from measurement error at 95% confidence. (A) Standard error of measurement (SEM, mm) by modality and timepoint. US demonstrated lower SEM than clinical photography at both timepoints: pre-operative SEM 0.058 mm (US) vs. 0.128 mm (clinical); post-operative SEM 0.049 mm (US) vs. 0.096 mm (clinical). (B) MDC (mm) by modality and timepoint. Pre-operative MDC: 0.160 mm (US) vs. 0.354 mm (clinical); post-operative MDC: 0.136 mm (US) vs. 0.267 mm (clinical). (C) MDC values for each modality and timepoint plotted against the mean observed surgical alveolar bone level change (US: 0.813 mm; Clinical: 0.679 mm). The orange dashed horizontal line represents a 0.5 mm a priori clinical significance threshold. All four MDC values fall well below the observed mean surgical change, confirming that both methods possessed sufficient precision to detect the magnitude of osseous change produced by the surgical procedures in this cohort. The mean surgical change exceeded the US MDC by a factor of 5.1× and the clinical MDC by 1.9×, indicating a substantially greater signal-to-noise ratio for ultrasound relative to clinical photography. SEM = standard error of measurement; MDC = minimum detectable change; US = ultrasound.

## DISCUSSION

The existing literature on intraoral ultrasonography for periodontal assessment has largely focused on cross-sectional studies that assess US measurement agreement with clinical probing or radiographic reference methods at a single time point. The studies have established that high-frequency intraoral US can accurately identify key anatomical landmarks, including the gingival margin, cementoenamel junction, and alveolar bone crest, and that US measurements agree reasonably well with conventional methods.[13, 14] What remained unaddressed was the more clinically consequential question: can intraoral US detect changes in alveolar bone level with sufficient precision to serve as a longitudinal monitoring tool? Our findings demonstrate that the US has a superb detection limit and can be used to monitor minute changes in alveolar bone levels that cannot be achieved with current clinical or radiographic protocols. This study is, to our knowledge, the first to evaluate US validity and measurement precision specifically in a longitudinal, within-patient, change-detection context using a concurrent clinical reference standard in human subjects undergoing a controlled surgical procedure that produces measurable bone-level change. Notably, a recently published paper from our team on US assessment of bone-level changes in an ex vivo animal model reached essentially the same conclusion.[12]

Chifor et al. used a 40 MHz device to measure gingival height and width before and 7 days after scaling in 18 teeth, demonstrating that US could detect post-scaling resolution of gingival edema [20]. That study, while informative, was limited to soft-tissue dimensions at a short follow-up interval. While the study showed that US could be used to monitor soft tissue changes, the study design does not permit a formal precision estimate using replicates.

The preoperative agreement between US and clinical alveolar bone level change measurements was excellent, with a Pearson correlation of r = 0.977 and a mean Bland-Altman bias of only −0.009 mm. The tight limits of agreement at baseline (±0.40 mm) are comparable to those reported in prior validation studies of intraoral US against conventional clinical probing. Figueredo et al. demonstrated excellent interrater ICC values for US-derived alveolar bone crest-to-CEJ distances (ICC 0.872–0.958) using a 20 MHz handheld transducer across 64 patients [13], and a subsequent ex vivo cadaver study by the same group confirmed intra- and inter-examiner ICC of 0.997 when comparing US to microCT as a reference standard.[21]

Postoperatively, the between-method correlation remained strong (r = 0.912) and the mean bias stayed negligible at +0.013 mm, confirming that US did not develop a systematic directional offset following periodontal surgery. The widening of the post-operative limits of agreement warrants careful interpretation. The post-operative clinical photographs used as the reference standard were obtained after surgery and before suturing, capturing bone-level architecture at the time of surgical completion. In contrast, the US imaging was performed two to six weeks postoperatively, following wound closure, soft tissue healing, and the early phase of osseous remodeling. The two methods, therefore, did not image the same biological moment. Over this interval, the alveolar crest could undergo remodeling, altering the CEJ-to-ABC distance.

The moderate-to-strong correlation between US-detected and clinically-detected surgical change at the site level (r = 0.802) confirms directional concordance between modalities at the individual-tooth level. Sites with minimal planned bone removal, comprising teeth treated primarily by osteoplasty or conservative ostectomy, contributed to the near-zero and negative change values observed in the distribution tails, and these sites appropriately showed small or indeterminate change on both modalities. Their inclusion reflects clinical reality and does not represent measurement failure.

The US MDC demonstrated in the present study was 0.160 mm preoperatively and 0.136 mm postoperatively. Notably, similar magnitudes of detection limit for bone-level changes were found in an ex vivo pig jaw model published by our team. Here, we demonstrated that the detection limit for bone loss ranged from 77 µm (automated) to 113 µm (manual), with a minimum detectable bone loss of 138 µm[12]. Also, our group recently evaluated US precision for soft-tissue monitoring in a separate human study, measuring gingival height from the alveolar bone crest in triplicate on 16 control teeth from 9 patients at two time points, 6 weeks apart, when no tissue changes were expected. The detection limit under those conditions was 0.09 mm (unpublished data). These are not marginal improvements; they represent a qualitative shift in what is measurable. To appreciate why this matters for periodontal monitoring, these values must be placed alongside the detection thresholds of the modalities currently used in clinical practice.

Haffajee et al.[22], showed that the difference in replicated clinical probing carries a mean measurement standard deviation of 0.82 mm. The MDC₉₅ should be 1.61 mm (1.96 x 0.82). However, authors used 3 x 0.82 or 3 mm as the threshold to indicate significant changes in probing attachment. These relatively high thresholds pose a challenge to studying site-specific disease activity. Radiography performs better in absolute terms but remains fundamentally limited: Hausmann et al.[6] estimated the radiographic system error at 0.87 mm under optimally controlled conditions, concluding that apparent bone-level changes at or below this threshold cannot be attributed to true biology. The US MDC demonstrated here — 0.160 mm for bone-level change and 0.09 mm for soft tissue (unpublished data) — is approximately five-fold below the radiographic floor and more than ten-fold below the probing-based threshold. Unlike clinical probing, where variability arises from probe angulation, applied force, tissue inflammation, and examiner interpretation, US interrogates hard and soft tissue interfaces directly and reproducibly, decoupled from the operator-dependent sources of noise that constrain conventional tools.[22] These are not marginal differences; they represent a qualitative shift in what is measurable.

This precision gap has direct implications for how periodontal disease progression is studied and detected clinically. As reviewed by Greenstein and Caton[23], reported rates of site-specific periodontal disease activity varied widely across longitudinal studies, because investigators were forced to set detection thresholds high to control false positives. The result is a systematic blind spot: thresholds set to avoid false positives inevitably generate false negatives, and incremental early disease activity goes undetected and uncharacterized. The US MDC demonstrated in the present study falls within this previously undetectable range, suggesting that intraoral US could enable detection of disease progression at a sensitivity that current tools structurally cannot achieve. Whether this advantage translates to clinically actionable monitoring in patients with active periodontitis remains to be confirmed in prospective longitudinal studies. Beyond precision, serial radiographic monitoring carries a practical constraint that limits its utility as a high-frequency surveillance tool: cumulative ionizing radiation exposure. While individual periapical exposures are low, the requirement for repeated examinations at 6-to 12-month intervals over a patient’s lifetime raises legitimate concerns under radiation protection principles, particularly for younger patients and those requiring monitoring across multiple sites. Intraoral US carries no radiation burden, imposes no interappointment waiting period, and can, in principle, be performed at every clinical visit — thereby enabling substantially more granular longitudinal data than is achievable with radiography under any clinically reasonable monitoring schedule.

### Limitations

Several limitations of this study require acknowledgment. The sample of 10 patients and 28 tooth sites, while sufficient to characterize agreement and precision in this proof-of-concept context, limits the generalizability of the findings. Future studies with larger cohorts and stratified surgical indications will be needed to confirm that these precision estimates are stable across the range of clinical presentations encountered in periodontal practice.

The clinical reference standard in this study was intraoperative photography rather than CBCT. We selected this approach deliberately: intraoperative photography provides direct visualization of the surgical field at the moment of osseous adjustment — the actual biological event of interest — and is not subject to the geometric distortion or beam-hardening artifacts that complicate CBCT assessment of thin buccal bone. However, it is inherently a two-dimensional measurement, does not capture the full three-dimensional bone architecture, and introduces uncertainty from angulation despite software correction. A future study incorporating CBCT at both timepoints would provide a volumetric reference standard capable of characterizing buccal and lingual bone-level changes simultaneously.

### Conclusions

Intraoral high-frequency ultrasonography demonstrated strong concurrent validity with clinical photography for measuring the CEJ-to-ABC distance in patients undergoing crown lengthening and osseous surgery, with preoperative agreement comparable to previously reported reliability studies. Critically, the US demonstrated a measurement-precision advantage, with a preoperative MDC of 0.160 mm, which falls substantially below the radiographic minimum detectable change. This precision advantage, combined with the absence of ionizing radiation and the feasibility of serial measurement at routine clinical visits, positions intraoral US as a potentially transformative tool for early, non-invasive longitudinal monitoring of alveolar bone-level change in periodontal patients. Larger prospective studies in patients with active periodontitis are warranted to confirm whether this precision translates to clinically actionable detection of disease progression.

## Data Availability

All data in the present study are available upon reasonable request to the authors.

## Acknowledgment

This work was supported by the National Institute of Health R01DE031307 awarded to J.V.J.

